# C-reactive protein-to-albumin ratio as a Novel Prognostic Biomarker for Long-Term Mortality in Pericarditis: A Real-World Study

**DOI:** 10.1101/2025.11.11.25339867

**Authors:** Lingyu Mi, Ishan Lakhani, Sharen Lee, Wing Tak Wong, Gary Tse, Fang Fang

## Abstract

**Background:** Pericarditis is a heterogeneous inflammatory condition with variable clinical outcomes. Although traditional inflammatory biomarkers such as C-reactive protein (CRP) and erythrocyte sedimentation rate (ESR) are routinely used for diagnosis and monitoring, they do not fully capture the interplay between inflammation, hepatic synthetic function, and nutritional status. The CRP–to–albumin ratio (CAR), a composite index integrating these components, has shown prognostic value in several cardiovascular disorders. However, its significance in pericarditis remains unknown.

**Methods:** This was a real-world retrospective cohort study of adult patients hospitalized for pericarditis between January 1^st^, 2005 to December 31^st^, 2019 from a single tertiary centre. CAR was calculated as CRP (mg/L) divided by serum albumin (g/L) and categorized into quartiles. The primary outcome was all-cause mortality. Associations were examined using Cox proportional hazards models, restricted cubic splines (RCS), and segmented Cox regression.

**Results:** A total of 546 patients (mean age, 59.2±16.4 years; 56.8% men) were analyzed. During a median follow-up of 64 months, 239 deaths (43.8%) occurred. Higher CAR quartiles were associated with progressively increased mortality (log-rank P<0.001). In multivariable Cox models adjusting for demographics and comorbidities, each unit increase in CAR conferred a 5% higher mortality risk (hazard ratio [HR], 1.05; 95% confidence interval [CI], 1.01–1.10; P=0.016). Compared with the lowest quartile, adjusted HRs for mortality were 2.31 (95% CI, 1.53–3.50), 2.65 (95% CI, 1.78–3.94), and 2.39 (95% CI, 1.58–3.60) across quartiles 2–4 (P for trend <0.001). RCS and segmented Cox analyses demonstrated a nonlinear relationship with a threshold near CAR=0.33—below which mortality risk increased sharply and plateaued thereafter. Associations were consistent across age, sex, hypertension, and malignancy subgroups.

**Conclusions:** CAR independently predicted long-term all-cause mortality in patients hospitalized for pericarditis, exhibiting a nonlinear dose–response pattern. CAR represents a simple, inexpensive, and readily available biomarker that integrates inflammatory and nutritional status, offering incremental prognostic value beyond traditional risk factors.

## Introduction

Pericarditis, defined as inflammation of the pericardial sac surrounding the heart, is a clinically important condition with diverse causes ^1,2^ and variable outcomes^2^. Although most patients recover with timely treatment, a subset may experience recurrent episodes or develop constrictive pericarditis, posing risks of morbidity and mortality^3,4^. Identifying prognostic indicators to stratify risk and guide management remains an unmet clinical need^4^.

Standard inflammatory biomarkers such as C-reactive protein (CRP) and erythrocyte sedimentation rate (ESR) are routinely used in the diagnosis and monitoring of pericarditis^2,5,6^. However, these markers may not capture the complex interplay among systemic inflammation, nutritional status, and hepatic synthetic function inherent in severe inflammatory states^7^. In cardiovascular research, composite biomarkers that integrate inflammatory and/ or nutritional components, such as the neutrophil-lymphocyte ratio ^8^, have gained increasing attention for their prognostic value in various cardiovascular conditions.

The CRP-to-albumin ratio (CAR) has emerged as a promising composite biomarker reflecting both inflammatory burden and nutritional reserve^9^. CRP rises rapidly as an acute-phase reactant in response to inflammatory stimuli, whereas serum albumin, a negative acute-phase protein, decreases during inflammation or catabolic states^7^. By combining both, CAR may provide a more comprehensive measure of the patient’s pathophysiological state^10,11^.

Recent investigations have demonstrated the prognostic utility of CAR across multiple cardiovascular settings. Elevated CAR levels have been independently associated with long-term mortality in patients with acute coronary syndromes^12^ and with increased long-term mortality among patients with chronic heart failure^13^, demonstrating prognostic value beyond conventional inflammatory markers such as CRP or ESR. In large community-based cohorts such as the United Kingdom Biobank, higher CAR levels were also linked to incident cardiovascular disease and all-cause mortality^14^. Collectively, these findings support CAR as an integrative biomarker of systemic inflammation, metabolic stress, and nutritional status.

Despite the growing evidence for the prognostic value of CAR in diverse cardiovascular conditions, its specific relevance in pericarditis has not been established. To date, no study has systematically evaluated the association between baseline CAR and long-term outcomes in this population^15^. Given the expanding therapeutic landscape and increasing emphasis on individualized management, reliable prognostic markers are urgently needed to guide risk stratification and patient follow-up in pericarditis^2,16^.

Accordingly, this study aimed to evaluate the association between baseline CAR and long-term all-cause mortality among patients hospitalized for pericarditis at a tertiary centre from Hong Kong using a subset of a prior study by our team ^17^. We further explored the dose–response and threshold relationships using restricted cubic spline and segmented Cox regression models, and conducted prespecified subgroup analyses by age, sex, hypertension, and malignancy status, to evaluate the consistency of associations across clinically relevant subgroups.

## Methods

### Study Design and Population

This study used a subset of a previously conducted retrospective cohort study of adult patients hospitalized for pericarditis between Jaunary 1^st^, 2005 to December 31^st^, 2019 from a single tertiary centre ^17^. The requirement for informed consent was specifically waived because of the retrospective design and use of deidentified data. The study was part of wider investigations approved by the Chinese University of Hong Kong–New Territories East Cluster Clinical Research Ethics Committee (https://www.crec.cuhk.edu.hk/approved-study-database/?dbr=&dbt=&dbn=tse+gary) on the use of ECGs for risk stratification (Approval numbers: 2019.338, 2019.361, and 2019.422), which contributed to the Doctor of Medicine degree of Dr. Gary Tse (https://www.proquest.com/docview/3252768154/278E578A36174385PQ). A letter from the MD Subcommittee of the Graduate School, The Chinese University of Hong Kong, authorizing the use of these data was provided. The associated datasets are publicly available via ProQuest (https://www.proquest.com/docview/3252768154/278E578A36174385PQ/2?%20Theses&sourcetype=Dissertations%20). The clinical database has been validated and widely applied in epidemiologic and cardiovascular outcomes research for single centre ^18,19^ as well as multi-centre studies ^20^, including cross-cluster studies from the territory ^21^.

We identified all adult patients (aged ≥18 years) who were hospitalized with a primary diagnosis of pericarditis between January 1^st^, 2005, and December 31^st^, 2019 from a single tertiary centre. Pericarditis was defined according to the European Society of Cardiology (ESC) diagnostic criteria and identified using International Classification of Diseases, Tenth Revision (ICD-10) codes upon discharge. For patients with multiple hospitalizations, only the index admission was included for analysis. Patients were excluded if they had missing baseline measurements of CRP or serum albumin, or lacked survival outcome data.

### Data Collection

Baseline demographic characteristics, comorbidities and laboratory parameters were extracted from the electronic health record system. Comorbid conditions—including hypertension, diabetes mellitus, sudden cardiac death (SCD), ischemic heart disease (IHD), acute myocardial infarction (AMI), malignant arrhythmia, hypertension, diabetes mellitus, peripheral vascular disease (PVD), malignancy, stroke or transient ischemic attack (TIA), chronic kidney disease (CKD), malignancy, and chronic obstructive pulmonary disease (COPD)—were identified using ICD-9 diagnostic codes. The Charlson Comorbidity Index (CCI) was calculated for each patient to quantify baseline disease burden. Laboratory measurements obtained during the index hospitalization included CRP, serum albumin, complete blood count, renal function tests, serum electrolytes, lipid profile, prothrombin time (PT), activated partial thromboplastin time (APTT), and high-sensitivity troponin I (hs-TnI).

The CRP–to–albumin ratio (CAR) was calculated as CRP (mg/L) divided by serum albumin (g/L), and patients were categorized into quartiles according to its distribution in the cohort.

### Outcomes

The primary outcome was all-cause mortality. Survival status was ascertained from hospital records and cross-checked with data from the Hong Kong Death Registry. Follow-up time was calculated from the date of index admission to the date of death or the last available follow-up record.

### Statistical Analysis

Continuous variables were summarized as means ± standard deviations for approximately normally distributed data or as medians with interquartile ranges for skewed data. Group differences across CAR quartiles were evaluated using the one-way ANOVA or the Kruskal–Wallis test, as appropriate. Categorical variables were presented as counts (percentages) and compared using the chi-square test.

Survival probabilities were estimated using the Kaplan–Meier method, and differences between CAR quartiles were compared with the log-rank test. The association between CAR and all-cause mortality was examined using Cox proportional hazards regression, and the proportional-hazards assumption was verified with Schoenfeld residuals. Hazard ratios (HRs) and 95% confidence intervals (CIs) were estimated for each quartile, with the lowest quartile serving as the reference group. P for trend was derived by treating quartiles as an ordinal variable.

To account for potential confounding, three sequential models were constructed: Model 1: an unadjusted model; Model 2: adjusted for age and sex; and Model 3: a fully adjusted model including comorbidities such as SCD, IHD, AMI, malignant arrhythmia, hypertension, diabetes mellitus, PVD, malignancy, CKD, stroke or TIA and COPD.

To evaluate nonlinear and threshold associations, restricted cubic spline (RCS) analyses with three knots (at the 10th, 50th, and 90th percentiles of CAR) were fitted within the Cox model, using the median CAR value as the reference. Sensitivity analyses were performed using less-adjusted models to verify consistency of the spline-derived patterns. The inflection point obtained from the multivariable RCS curve was further assessed using segmented Cox regression, and the likelihood-ratio test was applied to compare linear versus piecewise model fits.

Prespecified subgroup analyses were performed according to age (<65 vs ≥65 years), sex, hypertension, and malignancy status. These subgroups were selected based on prior literature and biological plausibility, given their potential to modify the relationship between systemic inflammation, nutritional status, and mortality^22–24^. Interaction terms between CAR and each subgroup variable were included in the fully adjusted model to assess effect modification.

All statistical analyses were conducted using R software (version 4.3.1; R Foundation for Statistical Computing, Vienna, Austria). A two-sided p value < 0.05 was considered statistically significant.

## Results

### Baseline Characteristics

A total of 546 patients hospitalized for pericarditis were included and stratified into quartiles of the CAR: Q1 (0.01–0.17), Q2 (0.17–0.95), Q3 (0.95–2.72), and Q4 (2.72–27.27) (**Figure 1**). The baseline characteristics across CAR quartiles are summarized in **Table 1**. Patients with higher CAR levels exhibited an overall worse clinical and biochemical profile. Specifically, elevated CAR was associated with higher heart rate (P < 0.001), lower serum sodium levels (P < 0.001), and decreased lipid concentrations—including total cholesterol, triglycerides, and low-density lipoprotein cholesterol (all P ≤ 0.022). Prothrombin time also increased progressively across CAR quartiles (P < 0.001). In addition, the proportion of male patients was higher in Q4 than Q1 (62.8% vs 44.4%; P = 0.046).

**Figure 1.**
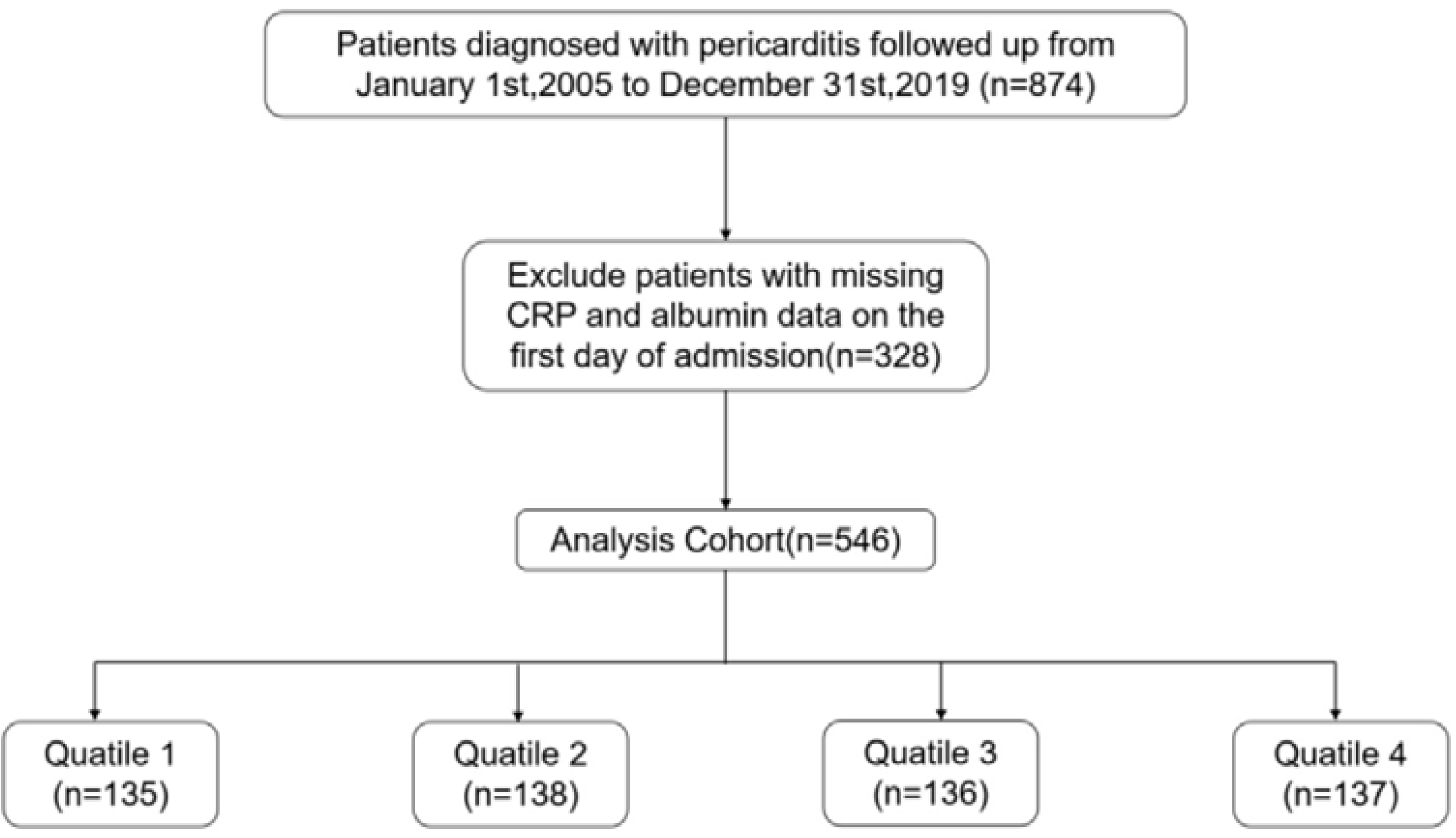
Flowchart of the study cohort. Flowchart illustrating patient inclusion and exclusion in the pericarditis cohort. **Abbreviations:** CAR indicates C-reactive protein–to–albumin ratio.

**Table 1.** Baseline Characteristics of Patients With Pericarditis According to Quartiles of the C-Reactive Protein–to–Albumin Ratio.

|  |  |  |  |  |  | P-value |
| --- | --- | --- | --- | --- | --- | --- |
| Categories | Overall(N= 546) | Q1 (N= 135) | Q2 (N= 138) | Q3 (N= 136) | Q4 (N= 137) | e |
| Age, years | 58.05 ± 17.35 | 60.39 ± 17.50 | 56.13 ± 19.34 | 57.69 ± 17.25 | 58.04 ± 14.97 | 0.242 |
| Gender |  |  |  |  |  | 0.046 |
| Female | 238 (43.59) | 60 (44.44) | 73 (52.90) | 54 (39.71) | 51 (37.23) |  |
| Male | 308 (56.41) | 75 (55.56) | 65 (47.10) | 82 (60.29) | 86 (62.77) |  |
| Heart rate,(beats/min) | 83.58 ± 21.70 | 74.42 ± 15.21 | 85.22 ± 21.97 | 87.96 ± 23.17 | 86.59 ± 22.92 | <.001 |
| Charlson | 2.55 ± 2.33 | 2.32 ± 1.59 | 2.77 ± 2.66 | 2.60 ± 2.47 | 2.49 ± 2.44 | 0.440 |
| Comorbidities |  |  |  |  |  |  |
| SCD, n(%) | 8 (1.47) | 2 (1.48) | 2 (1.45) | 1 (0.74) | 3 (2.19) | 0.881 |
| IHD, n(%) | 45 (8.24) | 14 (10.37) | 13 (9.42) | 7 (5.15) | 11 (8.03) | 0.425 |
| AMI, n(%) | 7 (1.28) | 2 (1.48) | 1 (0.72) | 1 (0.74) | 3 (2.19) | 0.686 |
| Malignant arrhythmia, n(%) | 8 (1.47) | 3 (2.22) | 4 (2.90) | 0 (0.00) | 1 (0.73) | 0.158 |
| Hypertension, n(%) | 90 (16.48) | 28 (20.74) | 22 (15.94) | 25 (18.38) | 15 (10.95) | 0.157 |
| DM, n(%) | 19 (3.48) | 4 (2.96) | 8 (5.80) | 1 (0.74) | 6 (4.38) | 0.096 |
| PVD, n(%) | 7 (1.28) | 2 (1.48) | 2 (1.45) | 2 (1.47) | 1 (0.73) | 0.921 |
| Malignancy, n(%) | 74 (13.55) | 15 (11.11) | 17 (12.32) | 24 (17.65) | 18 (13.14) | 0.418 |
| CKD, n(%) | 26 (4.76) | 5 (3.70) | 5 (3.62) | 8 (5.88) | 8 (5.84) | 0.693 |
| Stroke/TIA, n(%) | 17 (3.11) | 3 (2.22) | 7 (5.07) | 6 (4.41) | 1 (0.73) | 0.123 |
| COPD, n(%) | 5 (0.92) | 1 (0.74) | 2 (1.45) | 1 (0.74) | 1 (0.73) | 1.000 |
| Laboratory parameters |  |  |  |  |  |  |
| WBC, $\times 10^9/L$ | 7.75 ± 3.76 | 7.42 ± 3.07 | 7.13 ± 3.49 | 8.20 ± 4.13 | 8.35 ± 4.21 | 0.083 |
| RBC, $\times 10^{12}/L$ | 4.07 ± 0.83 | 4.23 ± 0.78 | 4.00 ± 0.72 | 3.95 ± 0.95 | 4.08 ± 0.85 | 0.126 |
| Platelet , $\times 10^9/L$ | 224.41 ± 98.51 | 222.12 ± 90.52 | 227.89 ± 96.14 | 223.33 ± 109.94 | 224.24 ± 99.24 | 0.982 |
| SMCH, pg | 30.20 ± 3.41 | 30.29 ± 3.43 | 29.98 ± 3.25 | 30.54 ± 3.67 | 29.98 ± 3.34 | 0.657 |
| Scr, $\mu\text{mol}/L$ | 84.00 (66.00, | 88.00 | 79.00 | 87.50 | 80.00 | 0.160 |
|  | 115.00) | (73.00,111.00) | (63.25,107.50) | (65.75,119.25) | (63.00,124.00) |  |
| Total cholesterol, $\text{mmol}/L$ | 3.94 ± 1.37 | 4.71 ± 1.53 | 3.68 ± 1.14 | 4.06 ± 1.23 | 3.39 ± 1.28 | <.001 |
| Triglycerides, $\text{mmol}/L$ | 1.01 (0.76, 1.43) | 1.05 (0.85,1.69) | 0.85 (0.70,1.24) | 1.05 (0.90,1.74) | 1.08 (0.79,1.61) | 0.022 |
| LDL-C, $\text{mmol}/L$ | 2.32 ± 1.07 | 2.85 ± 1.26 | 2.12 ± 0.98 | 2.37 ± 1.07 | 1.98 ± 0.74 | 0.006 |
| K$^+$, $\text{mmol}/L$ | 4.08 ± 0.59 | 4.12 ± 0.56 | 4.02 ± 0.56 | 4.04 ± 0.62 | 4.16 ± 0.62 | 0.164 |
| Na$^+$, $\text{mmol}/L$ | 137.76 ± 4.55 | 139.40 ± 3.68 | 138.21 ± 4.20 | 137.31 ± 4.53 | 136.13 ± 5.07 | <.001 |
| hs-TnI, $\text{ng}/L$ | 0.02 (0.01, 0.08) | 0.02 (0.01,0.06) | 0.02 (0.01,0.07) | 0.02 (0.01,0.06) | 0.04 (0.01,0.24) | 0.183 |
| APPT, second | 33.20 (30.30, 37.00) | 33.15 (29.95,36.42) | 33.30 (30.40,36.60) | 33.20 (30.05,37.50) | 33.30 (30.75,37.20) | 0.863 |
| PT, second | 12.00 (10.80, 13.70) | 11.00 (10.40,12.00) | 11.80 (10.70,13.60) | 12.60 (11.30,14.35) | 13.25 (11.43,14.70) | <.001 |

Data are expressed as mean ± SD, median (IQR), or number (%). Group differences were assessed using one-way ANOVA, Kruskal–Wallis, or $\chi^2$ tests, as appropriate.
**Abbreviations:** AMI indicates acute myocardial infarction; APTT, activated partial thromboplastin time; CAR, C-reactive protein–to–albumin ratio; CKD, chronic kidney disease; COPD, chronic obstructive pulmonary disease; DM, diabetes mellitus; hs-TnI, high-sensitivity troponin I; IHD, ischemic heart disease; IQR, interquartile range; LDL-C, low-density lipoprotein cholesterol; MCH, mean corpuscular hemoglobin; PT, prothrombin time; PVD, peripheral vascular disease; RBC, red blood cell; SCD, sudden cardiac death; Scr, serum creatinine; SD, standard deviation; TIA, transient ischemic attack; WBC, white blood cell.

### Kaplan–Meier Survival Analysis

During a median follow-up of 64 months, 239 deaths (43.8%) occurred. Kaplan–Meier survival curves revealed a graded, stepwise increase in all-cause mortality across CAR quartiles (log-rank P < 0.001) (**Figure 2**). Patients in the upper quartiles (Q3–Q4) showed markedly higher cumulative mortality, with early and sustained divergence of survival curves throughout follow-up.

**Figure 2.**
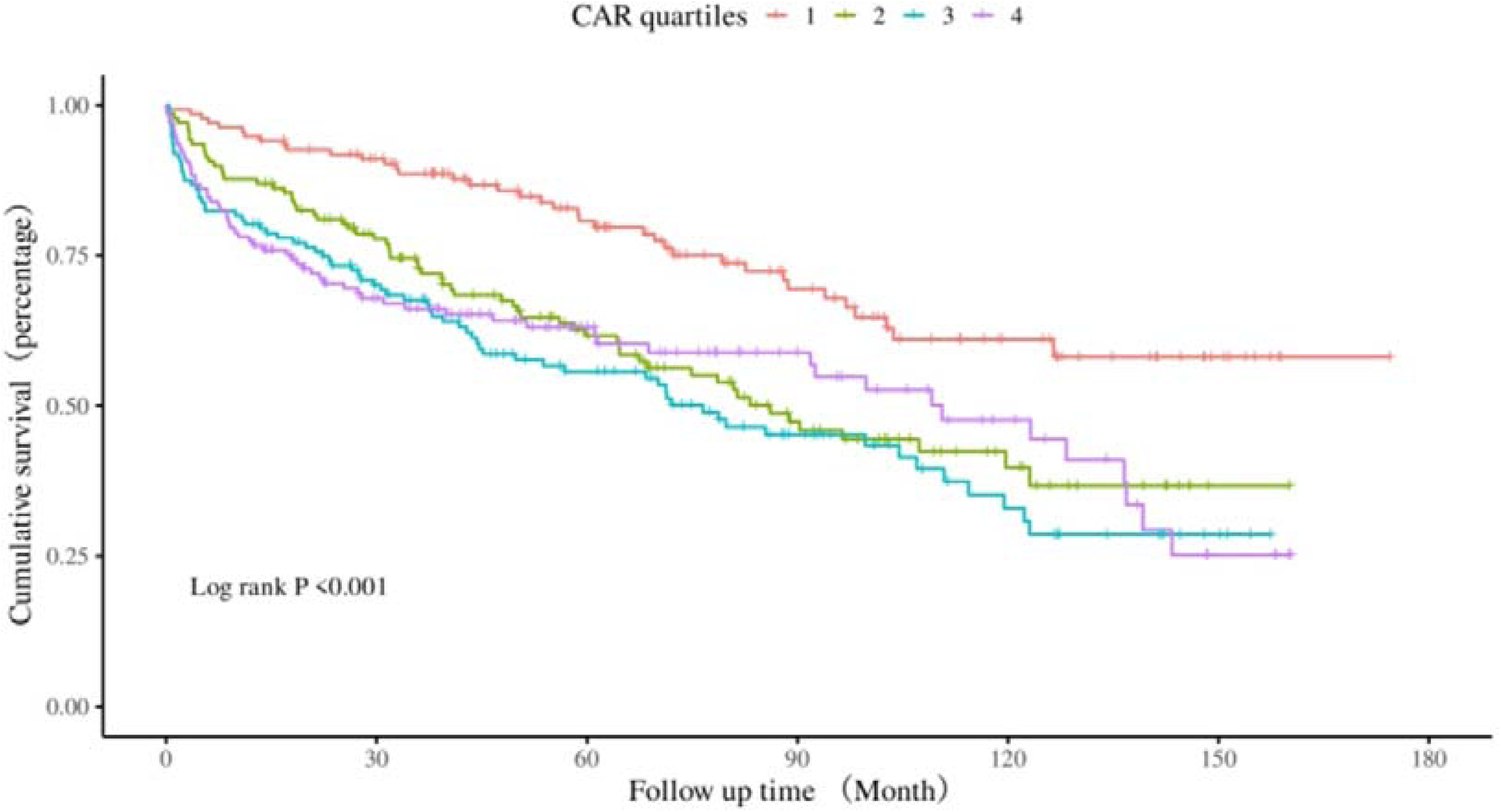
Kaplan–Meier Survival Curves for All-Cause Mortality According to Quartiles of the C-Reactive Protein–to–Albumin Ratio. Cumulative all-cause mortality curves across quartiles of the C-reactive protein–to–albumin ratio (Q1: 0.01–0.17; Q2: 0.17–0.95; Q3: 0.95–2.72; Q4: 2.72–27.27). Patients in higher quartiles showed progressively poorer survival (log-rank P < 0.001). Follow-up was measured from index admission to death or last contact. **Abbreviations:** CAR indicates C-reactive protein–to–albumin ratio.

### Multivariable Cox Regression Analysis

Multivariable Cox proportional hazards regression was performed to identify significant predictors (**Table 2**). In the fully adjusted model, CAR remained independently associated with all-cause mortality. When modeled as a continuous variable, each unit increase in CAR was associated with a 5% higher risk of death (HR, 1.05; 95% CI, 1.01–1.10; P = 0.016). When analyzed as quartiles, using the lowest quartile (Q1) as the reference, higher CAR categories were consistently associated with increased mortality risk. Adjusted HRs were 2.31 (95% CI, 1.53–3.50; P < 0.001) for Q2, 2.65 (95% CI, 1.78–3.94; P < 0.001) for Q3, and 2.39 (95% CI, 1.58–3.60; P < 0.001) for Q4 (P for trend < 0.001). These associations remained consistent across sequential adjustment models, indicating that the observed relationship was not driven by demographic or comorbidity confounding.

**Table 2.**
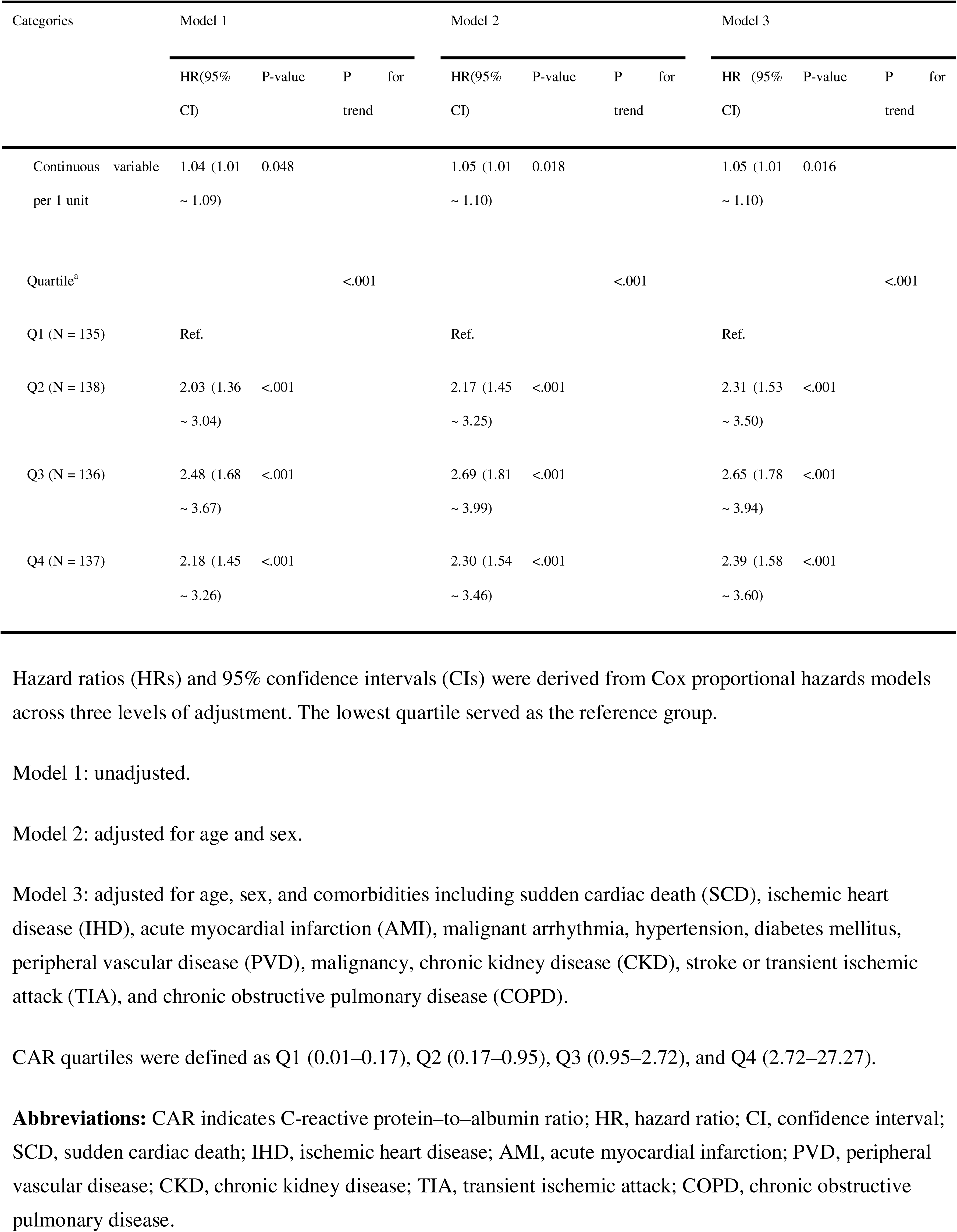
Cox Proportional Hazard Ratios for All-Cause Mortality According to Quartiles and Continuous Values of the C-Reactive Protein–to–Albumin Ratio.

| Categories |  | Model 1 |  |  |  | Model 2 |  |  |  | Model 3 |  |  |  |
| --- | --- | --- | --- | --- | --- | --- | --- | --- | --- | --- | --- | --- | --- |
|  |  | HR(95% | P-value | P | for | HR(95% | P-value | P | for | HR (95% | P-value | P | for |
|  |  | CI) |  | trend |  | CI) |  | trend |  | CI) |  | trend |  |
| Continuous | variable | 1.04 (1.01 | 0.048 |  |  | 1.05 (1.01 | 0.018 |  |  | 1.05 (1.01 | 0.016 |  |  |
|  | per 1 unit | ~ 1.09) |  |  |  | ~ 1.10) |  |  |  | ~ 1.10) |  |  |  |
| Quartile<sup>a</sup> |  |  |  | <.001 |  |  |  | <.001 |  |  |  | <.001 |  |
| Q1 (N = 135) |  | Ref. |  |  |  | Ref. |  |  |  | Ref. |  |  |  |
| Q2 (N = 138) |  | 2.03 (1.36 | <.001 |  |  | 2.17 (1.45 | <.001 |  |  | 2.31 (1.53 | <.001 |  |  |
|  |  | ~ 3.04) |  |  |  | ~ 3.25) |  |  |  | ~ 3.50) |  |  |  |
| Q3 (N = 136) |  | 2.48 (1.68 | <.001 |  |  | 2.69 (1.81 | <.001 |  |  | 2.65 (1.78 | <.001 |  |  |
|  |  | ~ 3.67) |  |  |  | ~ 3.99) |  |  |  | ~ 3.94) |  |  |  |
| Q4 (N = 137) |  | 2.18 (1.45 | <.001 |  |  | 2.30 (1.54 | <.001 |  |  | 2.39 (1.58 | <.001 |  |  |
|  |  | ~ 3.26) |  |  |  | ~ 3.46) |  |  |  | ~ 3.60) |  |  |  |

Hazard ratios (HRs) and 95% confidence intervals (CIs) were derived from Cox proportional hazards models across three levels of adjustment. The lowest quartile served as the reference group.
Model 1: unadjusted.
Model 2: adjusted for age and sex.
Model 3: adjusted for age, sex, and comorbidities including sudden cardiac death (SCD), ischemic heart disease (IHD), acute myocardial infarction (AMI), malignant arrhythmia, hypertension, diabetes mellitus, peripheral vascular disease (PVD), malignancy, chronic kidney disease (CKD), stroke or transient ischemic attack (TIA), and chronic obstructive pulmonary disease (COPD).
CAR quartiles were defined as Q1 (0.01–0.17), Q2 (0.17–0.95), Q3 (0.95–2.72), and Q4 (2.72–27.27).
**Abbreviations:** CAR indicates C-reactive protein–to–albumin ratio; HR, hazard ratio; CI, confidence interval; SCD, sudden cardiac death; IHD, ischemic heart disease; AMI, acute myocardial infarction; PVD, peripheral vascular disease; CKD, chronic kidney disease; TIA, transient ischemic attack; COPD, chronic obstructive pulmonary disease.

### Dose–Response Relationship

RCS analysis demonstrated a significant overall association between CAR and all-cause mortality (P overall < 0.001) and clear nonlinearity (P nonlinearity = 0.001) (**Figure 3**). The curve revealed a steep risk increase at lower CAR levels that gradually flattened at higher ranges.

**Figure 3.**
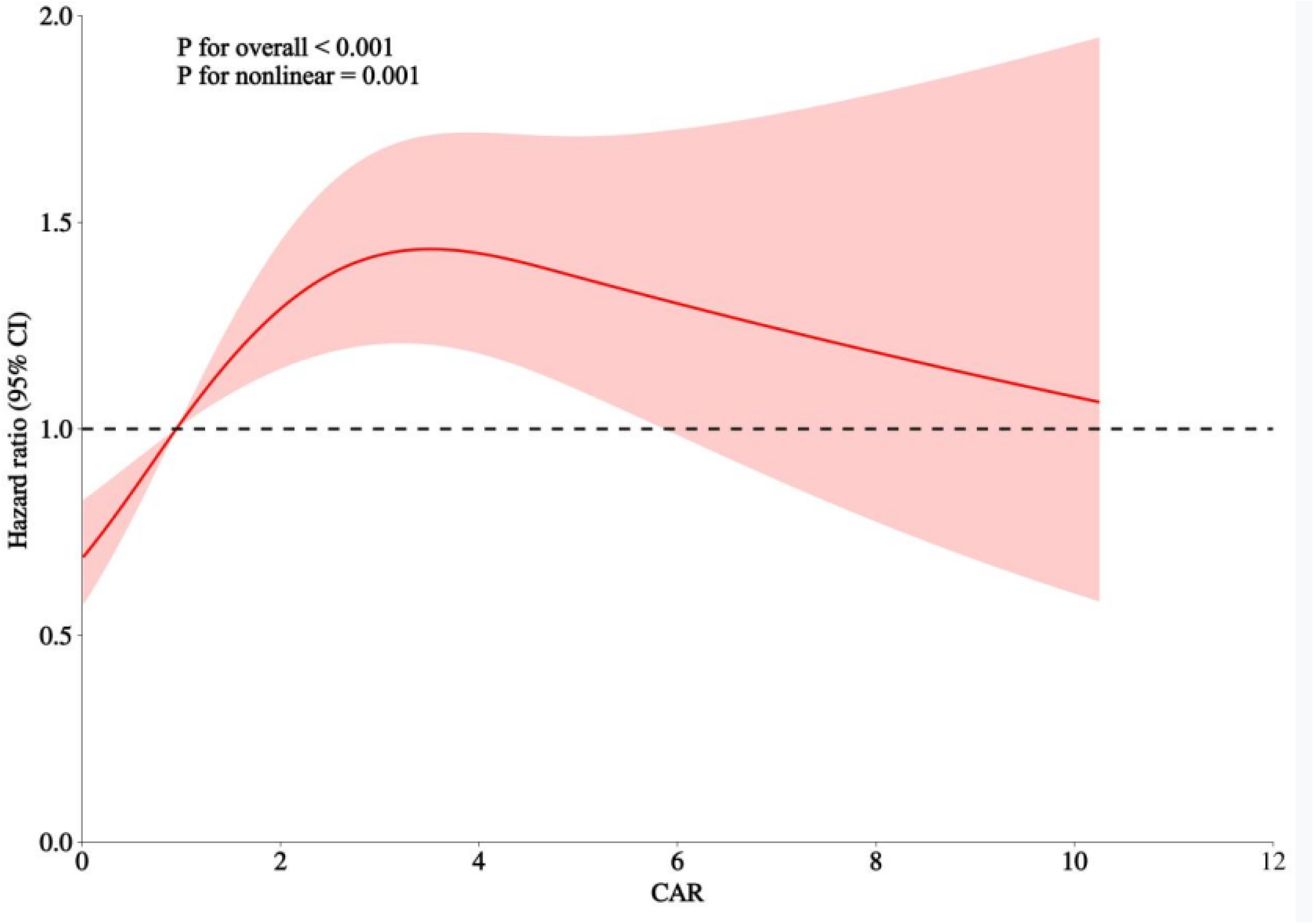
Multivariable-Adjusted Restricted Cubic Spline Analysis of the Association Between the C-Reactive Protein–to–Albumin Ratio and All-Cause Mortality. Restricted cubic spline analysis illustrating the nonlinear association between the C-reactive protein–to–albumin ratio (CAR) and all-cause mortality. Models were adjusted for age, sex, and comorbidities as in Model 3. The solid line represents the adjusted hazard ratio, and the shaded area indicates the 95% confidence interval. Knots were placed at the 10th, 50th, and 90th percentiles of CAR (P<0.001 for overall association; P=0.001 for nonlinearity). **Abbreviations:** CAR indicates C-reactive protein–to–albumin ratio; HR, hazard ratio; CI, confidence interval.

To further evaluate threshold effects, segmented Cox regression was performed (**Figure 4**), identifying an inflection point at a CAR value of approximately 0.33. Below this threshold, mortality risk increased sharply (adjusted HR: 24.88; 95% CI, 2.14–289.03; P = 0.010), although the wide confidence intervals reflect limited events in this range. Above this level, the association plateaued (adjusted HR: 0.99; 95% CI, 0.94–1.04; P = 0.689). The piecewise model provided a significantly better fit than a single linear term (likelihood-ratio P < 0.001) (**Supplementary Table S1**).

**Figure 4.**
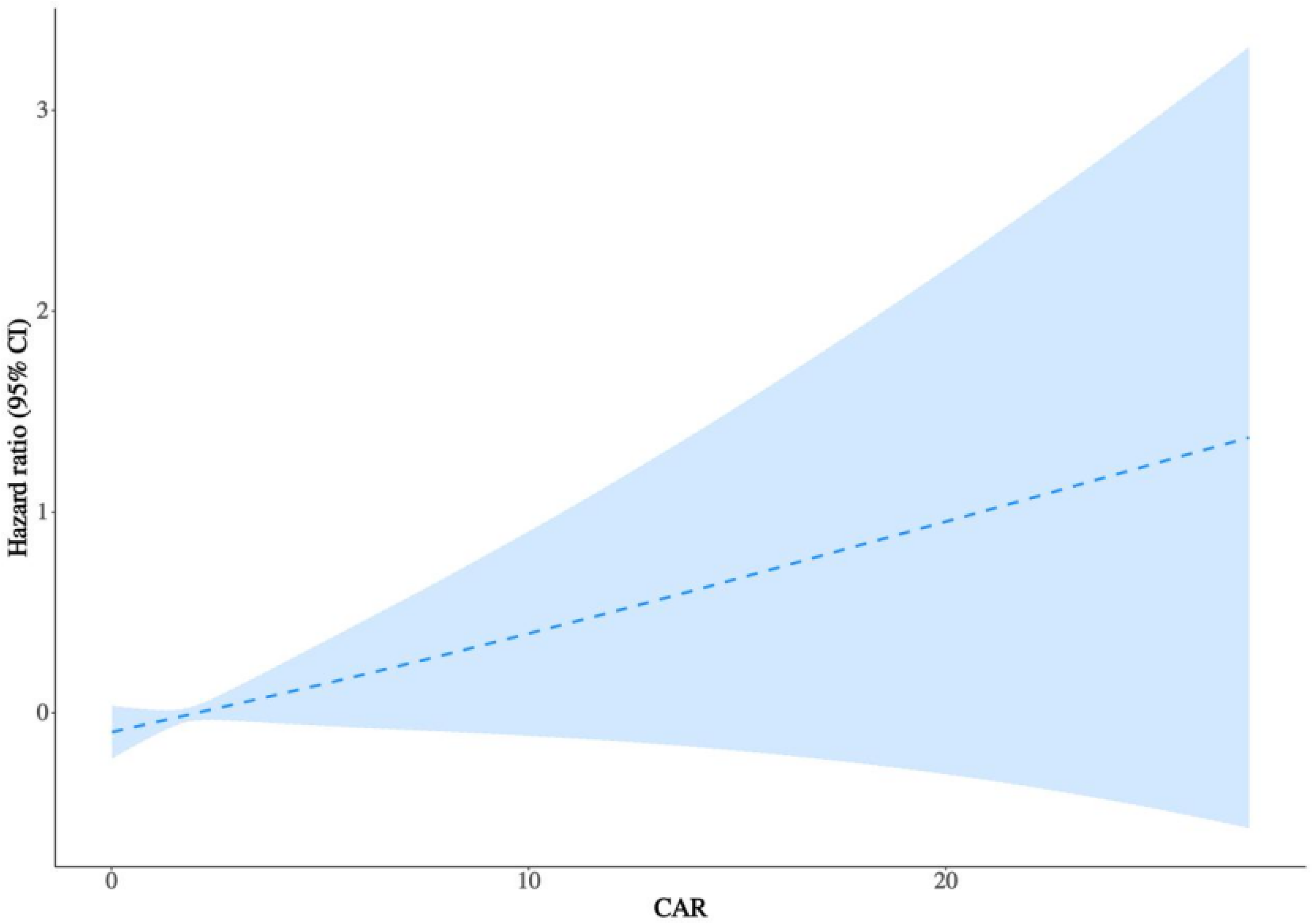
Segmented Cox Regression Illustrating the Threshold Effect of the C-Reactive Protein–to–Albumin Ratio on All-Cause Mortality. Segmented Cox regression illustrating the threshold effect of the C-reactive protein–to–albumin ratio (CAR) on all-cause mortality. The solid line represents the adjusted hazard ratio, and the shaded area indicates the 95% confidence interval. Models were adjusted for age, sex, and comorbidities as in Model 3. Below the threshold (CAR = 0.33), mortality risk increased steeply with CAR, whereas above this value the curve plateaued, suggesting a saturation effect (P<0.001 for likelihood-ratio test). **Abbreviations:** CAR indicates C-reactive protein–to–albumin ratio; HR, hazard ratio; CI, confidence interval.

### Sensitivity and Subgroup Analyses

Sensitivity analyses demonstrated that the association between CAR and all-cause mortality remained consistent across alternative model specifications. Restricted cubic spline analyses based on unadjusted, age- and sex-adjusted, and fully adjusted models (**Supplementary Figure S1**) revealed similar dose–response patterns, confirming the robustness of the observed nonlinear relationship.

In prespecified subgroup analyses (**Figure 5 and Table S2**), the positive association between higher CAR quartiles and all-cause mortality remained consistent across subgroups defined by sex, age, hypertension, and malignancy status. No significant interactions were observed (all P for interaction > 0.05), although the association appeared more pronounced among patients <65 years and those without malignancy.

**Figure 5.**
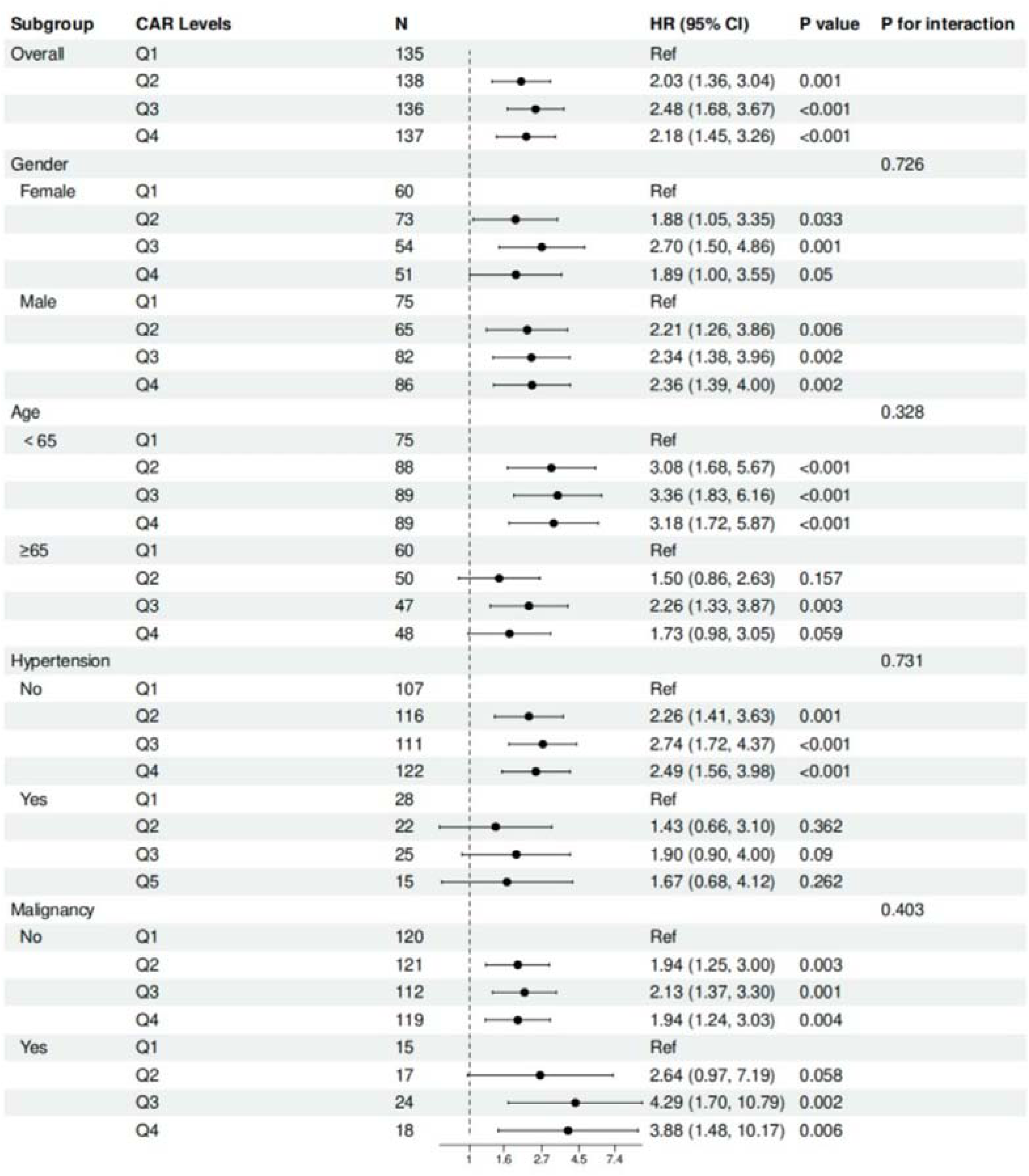
Subgroup Analysis of the Association Between the C-Reactive Protein–to–Albumin Ratio and All-Cause Mortality. Forest plot showing hazard ratios (HRs) and 95% confidence intervals (CIs) for all-cause mortality across quartiles of the C-reactive protein–to–albumin ratio (CAR) in prespecified subgroups defined by age, sex, hypertension, and malignancy status. Multivariable Cox regression models were used to estimate hazard ratios within each subgroup, adjusting for demographic and clinical covariates. Q1 served as the reference quartile. P-interaction values indicate effect modification across subgroups. Wider CIs in certain subgroups reflect limited sample size and event counts. **Abbreviations:** CAR indicates C-reactive protein–to–albumin ratio; HR, hazard ratio; CI, confidence interval.

## Discussion

### Principal Findings

In this cohort of patients hospitalized for pericarditis, CAR emerged as an independent predictor of long-term all-cause mortality. This association remained robust after multivariable adjustment for demographic and clinical covariates, highlighting the incremental prognostic utility of CAR beyond conventional clinical and inflammatory markers.

Restricted cubic spline and segmented Cox analyses revealed a nonlinear dose–response relationship, characterized by a steep rise in mortality risk at lower CAR values followed by a plateau at higher ranges, with a potential threshold at a CAR value of approximately 0.33. Although the hazard ratio below this threshold was markedly elevated, the wide confidence interval suggests limited precision, likely owing to the small number of events; nevertheless, the overall pattern supports a threshold effect.

These associations were consistent across prespecified subgroups (age, sex, hypertension, and malignancy status) and persisted in sensitivity analyses using alternative adjustment models, supporting the stability and robustness of the findings.

### Comparison with Previous Studies

Our findings are consistent with prior evidence underscoring the prognostic significance of CAR across a broad spectrum of cardiovascular disorders. In patients with acute coronary syndromes, elevated CAR has been independently associated with higher long-term mortality and adverse cardiac events^12^. Among coronary populations undergoing percutaneous coronary intervention, incorporation of CAR into predictive models further improved the discrimination of all-cause and cardiovascular mortality^25^.

Besides IHD, CAR has demonstrated prognostic relevance in other critical cardiovascular settings. Higher CAR levels were independently related to both in-hospital and six-month mortality in survivors of out-of-hospital cardiac arrest^26^. In heart-failure populations, elevated CAR correlated with worse functional profiles—such as higher New York Heart Association (NYHA) class—and was associated with increased in-hospital and out-of-hospital mortality^27^. In cohorts with heart failure and reduced ejection fraction, CAR independently predicted long-term mortality^28^.

Collectively, these studies identify CAR as a robust, integrative biomarker reflecting systemic inflammation and nutritional reserve—two interrelated processes central to cardiovascular pathophysiology. However, its prognostic utility may vary by clinical context. In a large community-based analysis from the UK Biobank, CAR did not outperform CRP in predicting incident cardiovascular events or mortality among otherwise healthy individuals, suggesting that its predictive value may be more pronounced in populations characterized by active inflammation or metabolic stress ^14^.

Our study extends this body of evidence by demonstrating a nonlinear association between CAR and mortality in pericarditis, with an inflection point near CAR = 0.33, below which mortality risk increased steeply before reaching a plateau. To our knowledge, few prior studies have examined such nonlinear or threshold effects in CAR-outcome relationships, highlighting the novelty and potential clinical relevance of our findings in this population.

### Pathophysiological Mechanisms

The prognostic relevance of CAR in pericarditis likely reflects the integrated effects of systemic inflammation, hepatic synthetic function, and nutritional reserve. Elevated CRP levels signify activation of innate immune pathways and the intensity of systemic inflammation^29,30^. Proinflammatory cytokines such as interleukin-1β, interleukin-6, and tumor necrosis factor-α stimulate hepatic CRP synthesis through NF-κB–dependent signaling. This cytokine-driven activation amplifies downstream inflammatory cascades, promoting pericardial injury, effusion, and fibrosis^31,32^. Consistent with these mechanisms, higher CRP levels have been associated with disease recurrence and an increased risk of constrictive remodeling in pericarditis^33,34^.

Conversely, hypoalbuminemia arises from cytokine-mediated suppression of hepatic synthesis^35^, increased vascular permeability^35^, and catabolic nutritional imbalance during acute systemic stress^36^. Low albumin not only indicates impaired nutritional and synthetic capacity but also contributes to oxidative stress and reduced plasma oncotic pressure^36^, both of which can exacerbate myocardial and pericardial inflammation^37^. Thus, an elevated CAR simultaneously captures heightened inflammatory burden and diminished homeostatic reserve—two interdependent processes that jointly drive adverse cardiovascular outcomes.

Unlike atherosclerotic or myocardial diseases, pericardial inflammation is characterized by an exudative cytokine milieu and fibrinous reaction within the pericardial cavity^2^. In this setting, hypoalbuminemia may further aggravate effusive-constrictive physiology by reducing plasma oncotic pressure, promoting pericardial effusion, and perpetuating the inflammatory–fibrotic cycle^38,39^. This mechanism provides a biological rationale linking elevated CAR to worse clinical outcomes in pericarditis^40^.

The observed CAR threshold near 0.33 may represent a biologically meaningful transition point. At lower CAR levels, even modest inflammation coupled with early albumin decline may trigger endothelial dysfunction and immune-metabolic activation, producing a disproportionate rise in mortality risk. Alternatively, elevated risk at the lowest CAR range may reflect chronic immune exhaustion or severe malnutrition, where suppressed inflammatory responses coexist with impaired physiological reserve, leading to vulnerability despite low apparent inflammation^40–42^. Beyond this range, further CAR elevation—largely driven by rising CRP levels— may reflect persistent inflammation without additional deterioration of systemic homeostasis, resulting in a plateau effect. This curvilinear pattern supports the concept that early disruption of inflammatory-nutritional equilibrium, rather than extreme inflammation alone, underlies the excess risk observed in pericarditis.

### Clinical Implications

Our findings support the clinical utility of CAR as a simple, inexpensive, and readily available biomarker for early risk stratification in pericarditis. Because it is derived from routinely measured laboratory parameters, CAR can be easily implemented across diverse clinical settings. In this study, a threshold near CAR = 0.33 emerged as a potential cutoff for identifying patients at disproportionately higher risk, suggesting that even modest elevations may carry important prognostic implications.

Importantly, CAR provides an objective and quantitative assessment of systemic inflammation and nutritional reserve, complementing established clinical risk factors such as fever, large pericardial effusion, and poor response to nonsteroidal anti-inflammatory drugs. This framework aligns with risk-adapted management strategies endorsed by the European Society of Cardiology and the American Heart Association/ American College of Cardiology, although current guidelines have yet to incorporate biomarker-based risk tools^3,43^.

Beyond baseline measurement, serial CAR monitoring may assist in tracking inflammatory activity, evaluating therapeutic response, and informing treatment adjustments—an approach consistent with emerging evidence supporting dynamic biomarker-guided follow-up in recurrent pericarditis^4,6^. In the longer term, individualized CAR-based risk profiles could enhance shared decision-making and improve communication between clinicians and patients regarding prognosis and care planning.

### Limitations

This study has several limitations. First, the retrospective observational design is inherently subject to residual confounding, despite comprehensive multivariable adjustment. Although major demographic and clinical covariates were accounted for, unmeasured factors including nutritional indices, proinflammatory cytokines, medication adherence, and comorbidity severity—could have influenced both CAR levels and outcomes^44^. Second, CAR was measured only at baseline, and temporal changes during hospitalization or follow-up were not captured. Thus, the prognostic significance of dynamic CAR trajectories remains unexplored. Previous evidence has suggested that dynamic inflammatory and nutritional responses may offer additional prognostic insight beyond baseline values^45^. Third, although this analysis was conducted on Chinese patients, the findings may not be fully generalizable to populations with different ethnic backgrounds, healthcare systems, or disease profiles. External validation in geographically and ethnically diverse cohorts is warranted to confirm the reproducibility of our results^46^. Finally, as with all retrospective studies, diagnostic accuracy and event classification depend on the completeness and quality of electronic records. Although segmented Cox regression identified a possible threshold effect around CAR = 0.33, the number of patients in the very low CAR range was limited, resulting in wide confidence intervals. Therefore, this exploratory finding should be interpreted cautiously and validated in larger, prospective multicenter studies.

## Conclusions

In this real-world cohort of patients hospitalized for pericarditis, the CAR was independently associated with long-term all-cause mortality. Both restricted cubic spline and segmented Cox regression analyses revealed a nonlinear relationship, with a threshold near CAR = 0.33, below which mortality risk increased steeply before reaching a plateau.

## Data Availability

This study was part of wider studies approved by the Chinese University of Hong Kong - New Territories East Cluster Clinical Research Ethics Committee (https://www.crec.cuhk.edu.hk/approved-study-database/?dbr=&dbt=&dbn=tse+gary) on the use of ECGs for risk stratification (Approval numbers: 2019.338, 2019.361, and 2019.422), which were used towards the Doctor of Medicine degree of Dr. Gary Tse (https://www.proquest.com/docview/3252768154/278E578A36174385PQ). A letter from the MD Subcommittee of the Graduate School, The Chinese University of Hong Kong, authorized the use of these data. The associated datasets are publicly available via ProQuest (https://www.proquest.com/docview/3252768154/278E578A36174385PQ/2?Theses&sourcetype=Dissertations).

## Acknowledgments

Not applicable.

## Supplementary Appendix

**Supplementary Figure S1.**
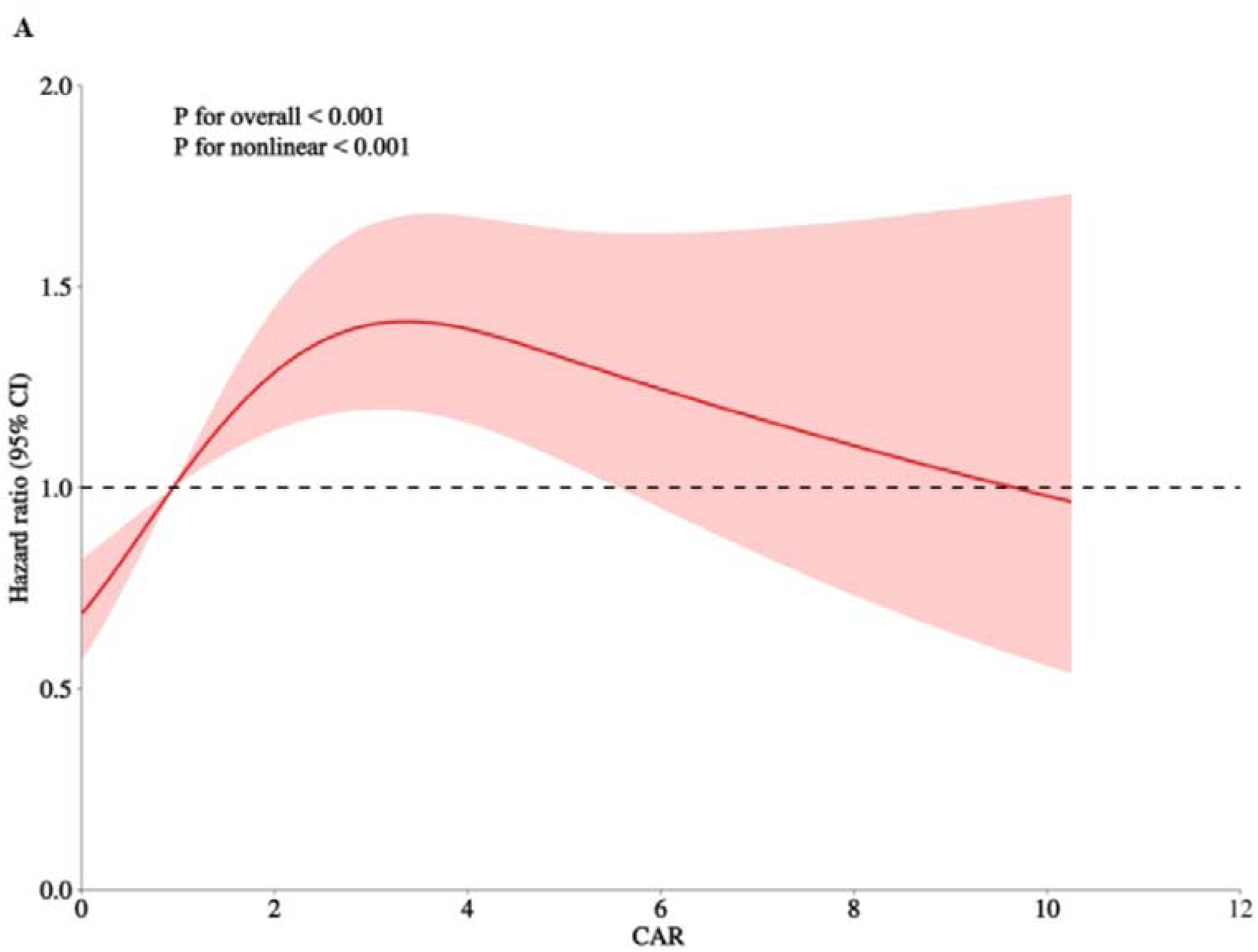
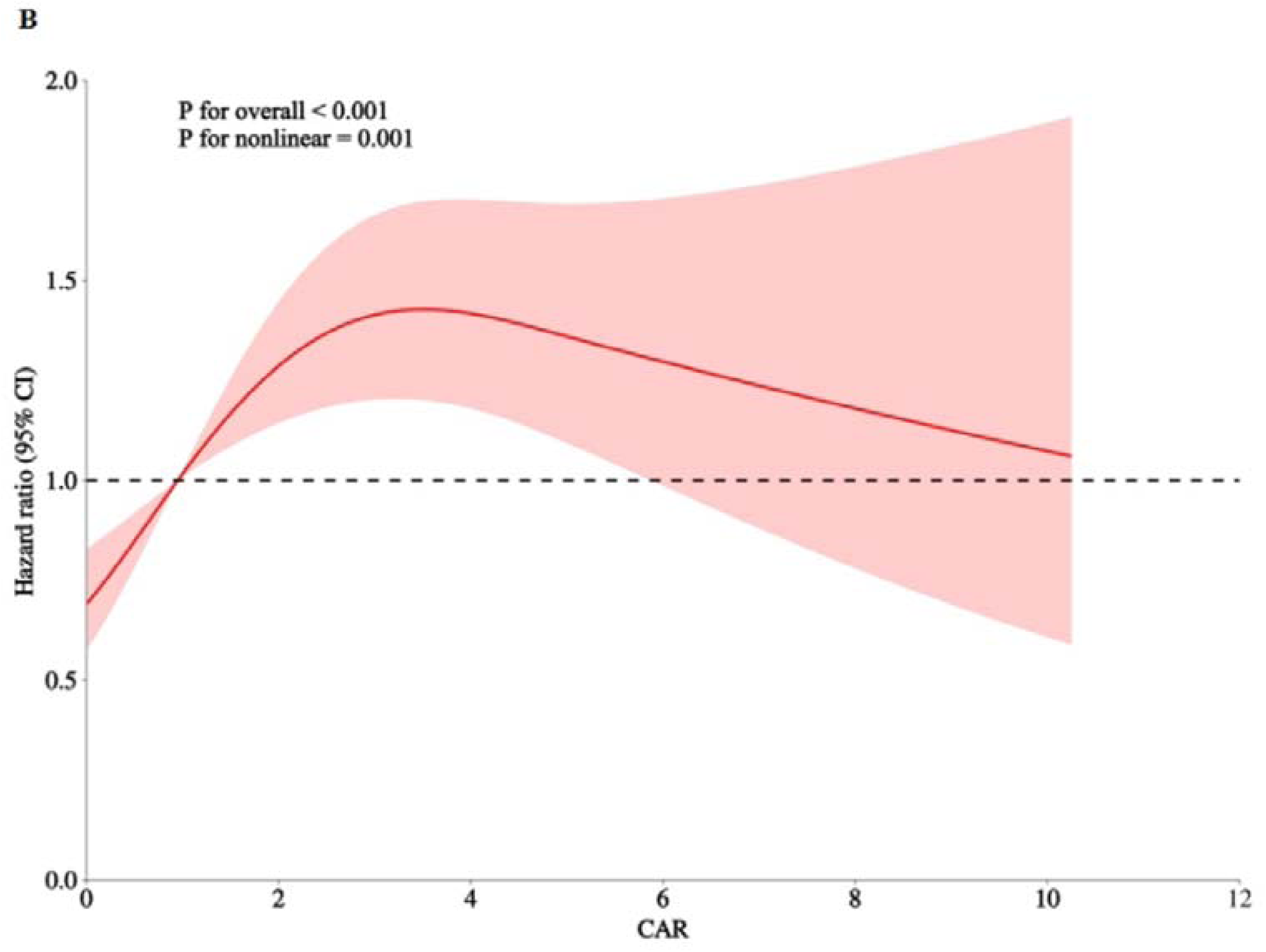
Restricted Cubic Spline Analyses of the Association Between the C-Reactive Protein–to–Albumin Ratio and All-Cause Mortality Conducted as Sensitivity Analyses. Restricted cubic spline (RCS) curves depicting the association between the C-reactive protein–to–albumin ratio (CAR) and all-cause mortality. (A) Unadjusted model. (B) Model adjusted for age and sex. The solid line represents the estimated hazard ratio, and the shaded area indicates the 95% CI. The dashed line denotes HR = 1.0. Knots were placed at the 10th, 50th, and 90th percentiles of CAR. Results of the fully adjusted model are presented in Figure 3 (P overall < 0.001; P nonlinearity < 0.001). **Abbreviations:** CAR indicates C-reactive protein–to–albumin ratio; HR, hazard ratio; CI, confidence interval.

**Supplementary Table S1.**
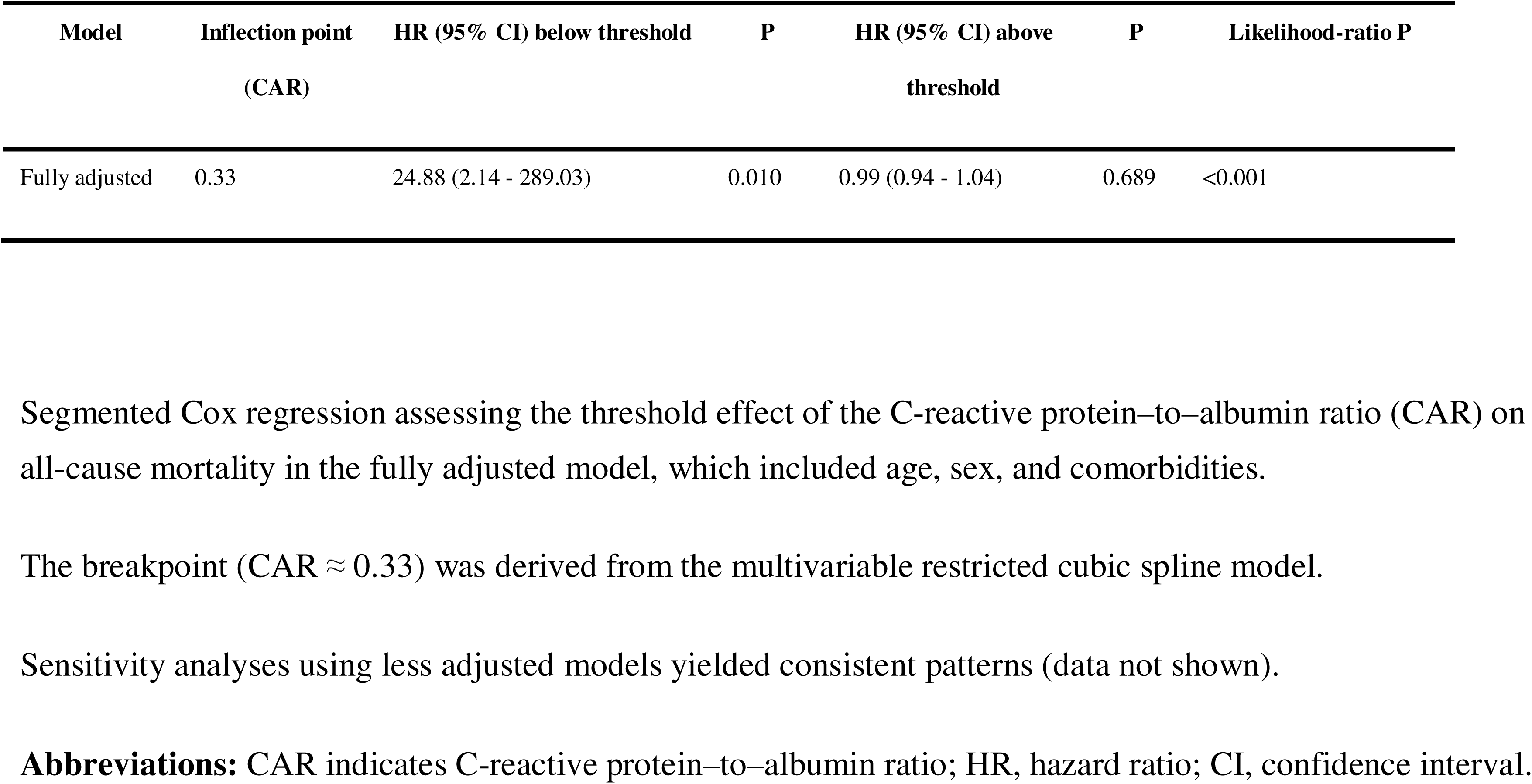
Threshold Effect of the C-Reactive Protein–to–Albumin Ratio on All-Cause Mortality in the Fully Adjusted Model.

**Supplementary Table S2.**
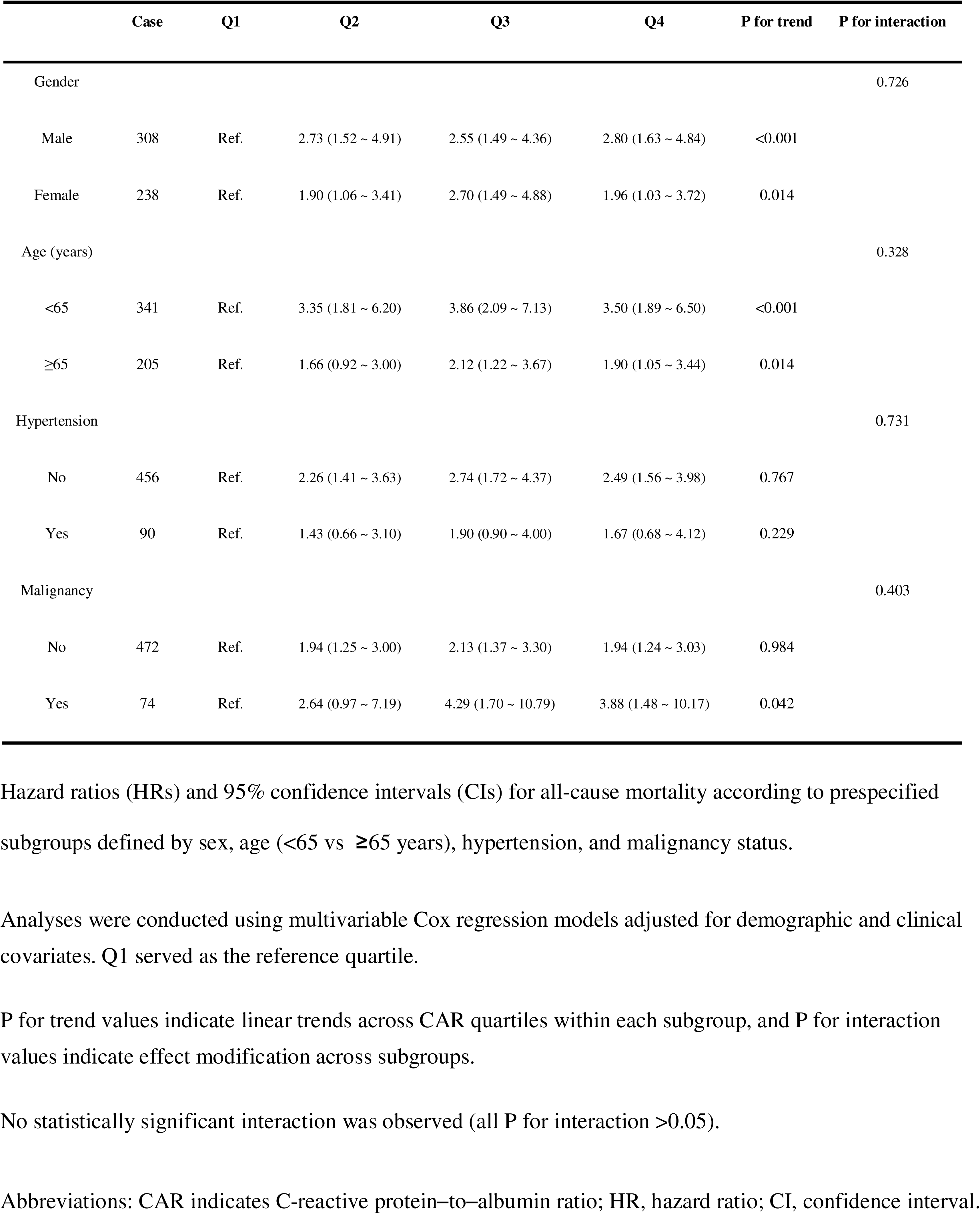
Subgroup Analyses of the Association Between the C-Reactive Protein–to–Albumin Ratio and All-Cause Mortality.

---

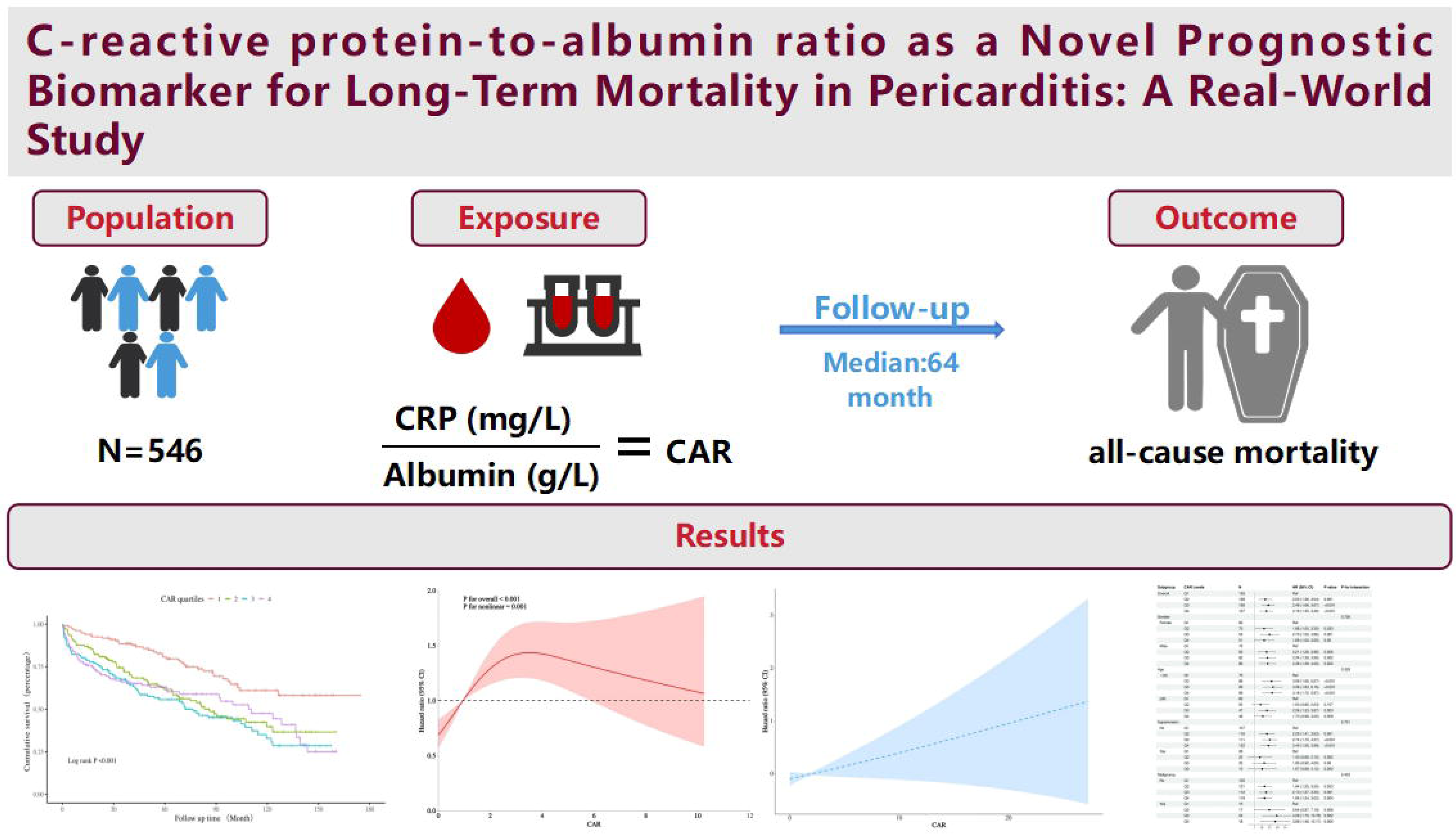

