## Supplemental Table S1and S2 and Figure S1 for "C-reactive protein-to-albumin ratio as a Novel Prognostic Biomarker for Long-Term Mortality in Pericarditis: A Real-World Study"

**Supplementary Figure S1.** Restricted Cubic Spline Analyses of the Association Between the C-Reactive Protein–to–Albumin Ratio and All-Cause Mortality Conducted as Sensitivity Analyses

A

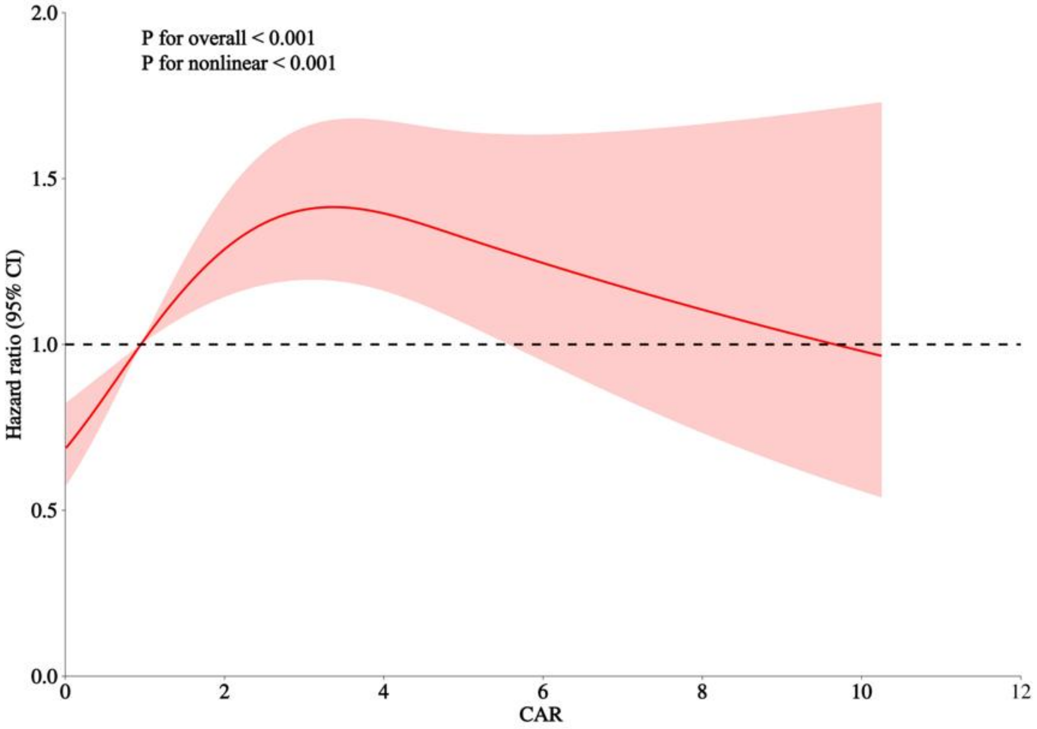

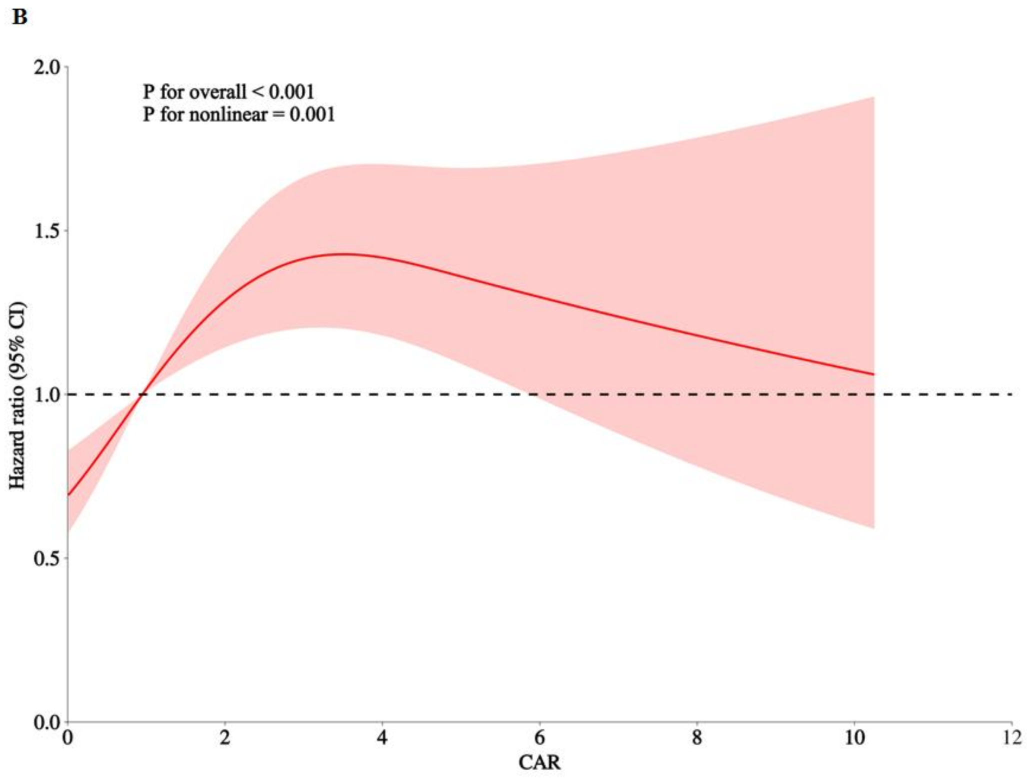


Restricted cubic spline (RCS) curves depicting the association between the C-reactive protein–to–albumin ratio (CAR) and all-cause mortality.

(A) Unadjusted model. (B) Model adjusted for age and sex. The solid line represents the estimated hazard ratio, and the shaded area indicates the 95% CI. The dashed line denotes HR = 1.0.

Knots were placed at the 10th, 50th, and 90th percentiles of CAR. Results of the fully adjusted model are presented in Figure 3 (P overall < 0.001; P nonlinear < 0.001).

**Abbreviations:** CAR indicates C-reactive protein–to–albumin ratio; HR, hazard ratio; CI, confidence interval.

**Supplementary Table S2.** Threshold Effect of the C-Reactive Protein–to–Albumin Ratio on All-Cause Mortality in the Fully Adjusted Model

| Model | Inflection point (CAR) | HR (95% CI) below threshold | P | HR (95% CI) above threshold | P | Likelihood-ratio P |
|---|---|---|---|---|---|---|
| Fully adjusted | 0.33 | 24.88 (2.14 - 289.03) | 0.010 | 0.99 (0.94 - 1.04) | 0.689 | <0.001 |

Segmented Cox regression assessing the threshold effect of the C-reactive protein–to–albumin ratio (CAR) on all-cause mortality in the fully adjusted model, which included age, sex, and comorbidities.

The breakpoint (CAR ≈ 0.33) was derived from the multivariable restricted cubic spline model.

Sensitivity analyses using less adjusted models yielded consistent patterns (data not shown).

**Abbreviations:** CAR indicates C-reactive protein–to–albumin ratio; HR, hazard ratio; CI, confidence interval.

**Supplementary Table S3.** Subgroup Analyses of the Association Between the C-Reactive Protein–to–Albumin Ratio and All-Cause Mortality

| | Case | Q1 | Q2 | Q3 | Q4 | P for trend | P for interaction |
|---|---|---|---|---|---|---|---|
| Gender | | | | | | | 0.726 |
| Male | 308 | Ref. | 2.73 (1.52 ~ 4.91) | 2.55 (1.49 ~ 4.36) | 2.80 (1.63 ~ 4.84) | <0.001 | |
| Female | 238 | Ref. | 1.90 (1.06 ~ 3.41) | 2.70 (1.49 ~ 4.88) | 1.96 (1.03 ~ 3.72) | 0.014 | |
| Age (years) | | | | | | | 0.328 |
| <65 | 341 | Ref. | 3.35 (1.81 ~ 6.20) | 3.86 (2.09 ~ 7.13) | 3.50 (1.89 ~ 6.50) | <0.001 | |
| ≥65 | 205 | Ref. | 1.66 (0.92 ~ 3.00) | 2.12 (1.22 ~ 3.67) | 1.90 (1.05 ~ 3.44) | 0.014 | |
| Hypertension | | | | | | | 0.731 |
| No | 456 | Ref. | 2.26 (1.41 ~ 3.63) | 2.74 (1.72 ~ 4.37) | 2.49 (1.56 ~ 3.98) | 0.767 | |
| Yes | 90 | Ref. | 1.43 (0.66 ~ 3.10) | 1.90 (0.90 ~ 4.00) | 1.67 (0.68 ~ 4.12) | 0.229 | |
| Malignancy | | | | | | | 0.403 |
| No | 472 | Ref. | 1.94 (1.25 ~ 3.00) | 2.13 (1.37 ~ 3.30) | 1.94 (1.24 ~ 3.03) | 0.984 | |
| Yes | 74 | Ref. | 2.64 (0.97 ~ 7.19) | 4.29 (1.70 ~ 10.79) | 3.88 (1.48 ~ 10.17) | 0.042 | |

Hazard ratios (HRs) and 95% confidence intervals (CIs) for all-cause mortality according to prespecified subgroups defined by sex, age (<65 vs ≥65 years), hypertension, and malignancy status.

Analyses were conducted using multivariable Cox regression models adjusted for demographic and clinical covariates. Q1 served as the reference quartile.

P for trend values indicate linear trends across CAR quartiles within each subgroup, and P for interaction values indicate effect modification across subgroups.

No statistically significant interaction was observed (all P for interaction >0.05).

**Abbreviations:** CAR indicates C-reactive protein–to–albumin ratio; HR, hazard ratio; CI, confidence interval.
